# A Prospective Controlled Study of Postpartum Breastfeeding Rates in Women with Type 2 Diabetes: Does Obesity Matter More Than Diabetes?

**DOI:** 10.64898/2026.01.18.26344373

**Authors:** Fiona L Britten, Emma L Duncan, Leonie K Callaway

**Affiliations:** Department of Obstetric Medicine, Women’s and Newborn Services, Royal Brisbane and Women’s Hospital, Herston, Brisbane, Queensland, Australia 4029; Department of Endocrinology, Royal Brisbane and Women’s Hospital, Brisbane, Queensland, Australia 4029; University of Queensland, Brisbane, Queensland 4006, Australia; Department of Twin Research & Genetic Epidemiology, School of Life Course Sciences, Faculty of Life Sciences and Medicine, King’s College London, London SE1 7EH, UK; Department of Endocrinology, Guy’s & St Thomas’ NHS Foundation Trust, London, UK

**Keywords:** type 2 diabetes, breastfeeding, lactation

## Abstract

Breastfeeding is associated with metabolic benefits for women with type 2 diabetes and their infants, yet breastfeeding rates in these women are poorly described. Obesity is associated with type 2 diabetes, and lower breastfeeding rates, making it difficult to disentangle type 2 diabetes vs. obesity effects on breastfeeding.

**Aims:** The primary objective was to examine breastfeeding rates in women with type 2 diabetes, compared to normoglycaemic women with either (1) matched body mass index (BMI) or (2) normal BMI. The secondary objective was to examine variables associated with breastfeeding.

**Methods:** Pregnant women with type 2 diabetes (cases) or normoglycaemia (controls) were prospectively recruited. Each case was matched with two controls by age and parity and either (1) matched or (2) normal (18–25 kg/m²) BMI. Data were collected from in-person surveys antenatally, medical records, and a four-month postpartum telephone survey. Analysis included descriptive and inferential statistics.

**Results:** Four-month breastfeeding data were available for 29, 29 and 28 women in each group. Breastfeeding rates were similar in women with type 2 diabetes and BMI-matched controls, both significantly lower than normal-BMI controls.

**Conclusions:** Maternal obesity, rather than type 2 diabetes, may be the major determinant of reduced breastfeeding rates four-months postpartum.

## 1.0 Introduction

Breastfeeding is associated with significant long term metabolic benefits for women with type 2 diabetes and their children, lowering risks of cardiovascular disease[1] and stroke in mothers[2]; and of diabetes in their infants[3]. Rates of breastfeeding in these women after hospital discharge are poorly described: some studies suggest women with type 2 diabetes have lower rates of breastfeeding than normoglycaemic women, although these studies are inconsistent[4, 5].

Obesity is associated with decreased breastfeeding rates; but it is unclear whether this is due to biological and/or social drivers, to increased peripartum medical interventions, also associated with lower breastfeeding rates[6]; or to other mechanisms. As many women with type 2 diabetes also have overweight or obesity, it is difficult to disentangle independent effects of type 2 diabetes and obesity, and any interactions between these variables, on breastfeeding rates.

The primary objective of this prospective study was to examine breastfeeding rates four months postpartum in women with type 2 diabetes compared to age- and parity-matched normoglycaemic women, with either (1) matched body mass index (BMI); or (2) normal-range BMI. The secondary objective was to examine variables in women with type 2 diabetes associated with breastfeeding four-months postpartum.

## 2.0 Materials and Methods

This study was approved by the Royal Brisbane and Women’s Hospital Ethics Committee (HREC/13QRBW/356) and all participants gave written informed consent.

### 2.1 Cohort Recruitment

Women with a pre-pregnancy diagnosis of type 2 diabetes were prospectively recruited from an obstetric endocrinology clinic, between May 2014 and December 2020. Inclusion criteria were: all eligible women aged ≥18 years, who could speak English, give informed consent, and intended to breastfeed. In addition to not fulfilling inclusion criteria, other exclusion criteria included history of prolactinoma.

Each case was matched 1:1 with an age-, parity- and pre-pregnancy BMI-matched normoglycaemic control termed “BMI-matched controls” and an age-, parity- and normal (18-25 kg/m^2^) pre-pregnancy BMI normoglycaemic control termed “normal-BMI controls,” prospectively recruited from general obstetric- or midwifery-led clinics. Controls were excluded if they had prolactinoma, other pregestational diabetes, impaired glucose tolerance, or gestational diabetes. Matching parameters were: age within five years, parity (nulliparous or multiparous), and prepregnancy BMI within five points (where relevant). Recruitment occurred in third trimester, after routine oral glucose tolerance testing (24-28 weeks’ gestation), to avoid inadvertent recruitment of control women with gestational diabetes.

### 2.2 Data collection

All data collection tools are presented in Supplementary Tables S2-S4.

Pre-partum maternal demographic data (collected by in-person survey and medical record review) included age, parity, pre-pregnancy BMI, medical history, diabetes medications, employment, income and breastfeeding intentions using the validated Infant Feeding Intentions (IFI) scale[7].

Peripartum maternal and infant variables (collected from the medical record) included mode of delivery, analgesia, neonatal hypoglycaemia, neonatal nursery admission, maternal/infant pregnancy/birth complications, and early feeding methods and frequency.

Breastfeeding data (collected at four months postpartum by telephone survey) included method of feeding. ‘Full’ breastfeeding was defined as breastfeeding or expressed breastmilk given as only milk feed used at time of survey regardless of previous formula use, and concurrent use of other non-milk fluids such as water, juice and medicine. Mixed feeding included breastmilk and formula/other fluids as outlined above. Formula feeding included formula as the only milk feed given. Reasons for formula use and timing and reason for breastfeeding cessation were recorded. The four month time point was chosen as it allowed sufficient time for breastfeeding to become well-established, but was before interfering factors (solid food initiation, maternal return-to-work) become common in Australian populations)[8, 9].

### 2.3 Power calculations

There are few data to inform power calculations for breastfeeding studies*. A priori*, a convenience sample enrolment of 60 women per group was planned. A study published during study recruitment suggested 20 women per group would give 90% power to detect a significant difference in ‘full’ breastfeeding based on a ‘full’ breastfeeding rate of 23% in women with type 2 diabetes at four months compared to 62% in normoglycaemic population controls[10]. The current study recruited 35 women to each group (type 2 diabetes, both control groups), with recruitment affected by the COVID-19 pandemic. The data evaluation presented in this paper shows a statistically significant difference in ‘full’ breastfeeding between the type 2 diabetes and normal-BMI control group, confirming sufficient recruitment for meaningful analysis.

### 2.4 Statistical methods

Baseline demographic, peripartum, and four-month postpartum outcomes were compared across groups using one-way ANOVA (mean ± standard error) for normally distributed data and Kruskal–Wallis tests (median [IQR]) for non-normal data, with Dunn’s correction where indicated. Categorical variables were analysed using chi-squared tests or Fisher’s exact tests for small cell counts. Analyses were conducted at the group level (not pairwise) due to dropout across all groups between data collection points. Post-hoc univariate analyses examined factors associated with ‘full’ and ‘any’ breastfeeding at four months among women with type 2 diabetes.

## 3.0 Results

Thirty-two women with type 2 diabetes, 33 BMI-matched controls, and 35 normal-BMI controls women completed the baseline antenatal survey. Subsequently, two participants declined further study participation due to traumatic birth and breastfeeding experience, and twelve were unable to be contacted postpartum. Four-month postpartum data were available for 29 of 32 (91%), 29 of 33 (88%) and 28 of 35 (80%) participants from each group respectively (Table 1). Considering matching of remaining participants: women with type 2 diabetes remained well-matched for age and parity to both control groups, and well-matched for BMI with BMI-matched controls. Women with type 2 diabetes and their matched BMI controls had significantly higher BMI (at both time points) compared with normal-range BMI controls (see Table 1). Women who discontinued the study prior to the postpartum survey had similar demographic and peripartum variables to women who completed the study although small numbers precluded inferential analysis (data not shown). All further analyses were based on women who provided both baseline and four-month postpartum breastfeeding data.

**Table 1.**
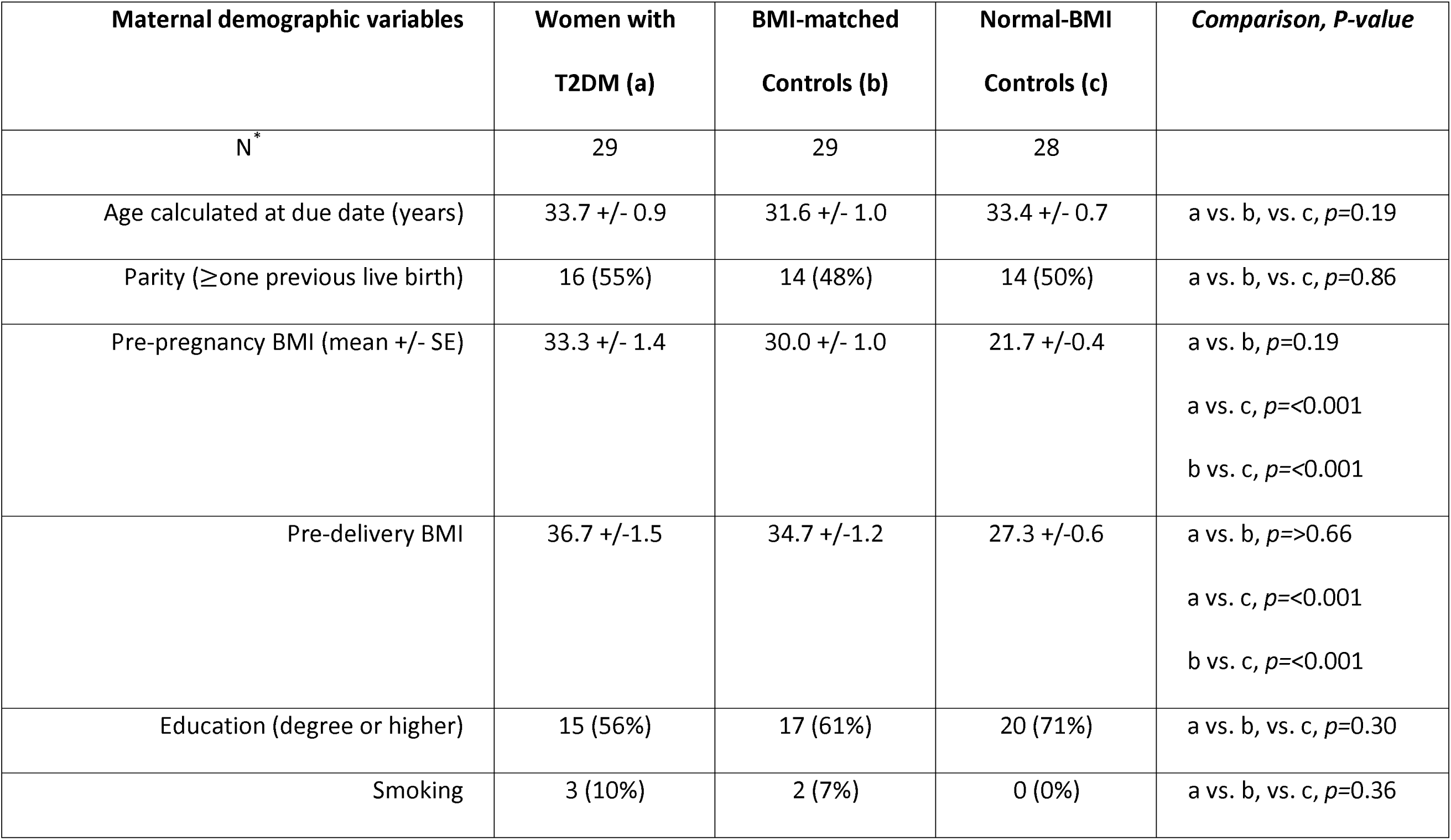
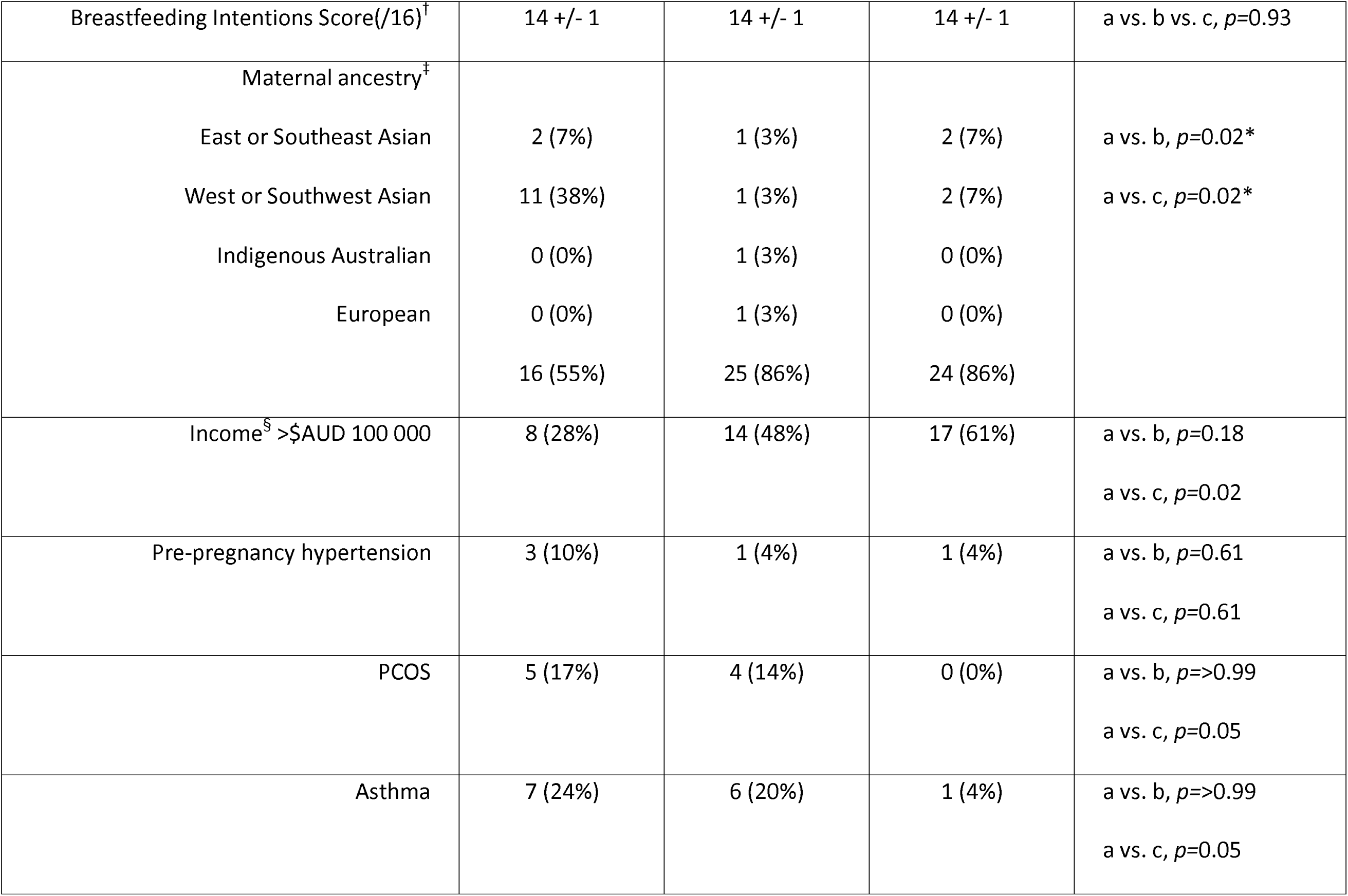
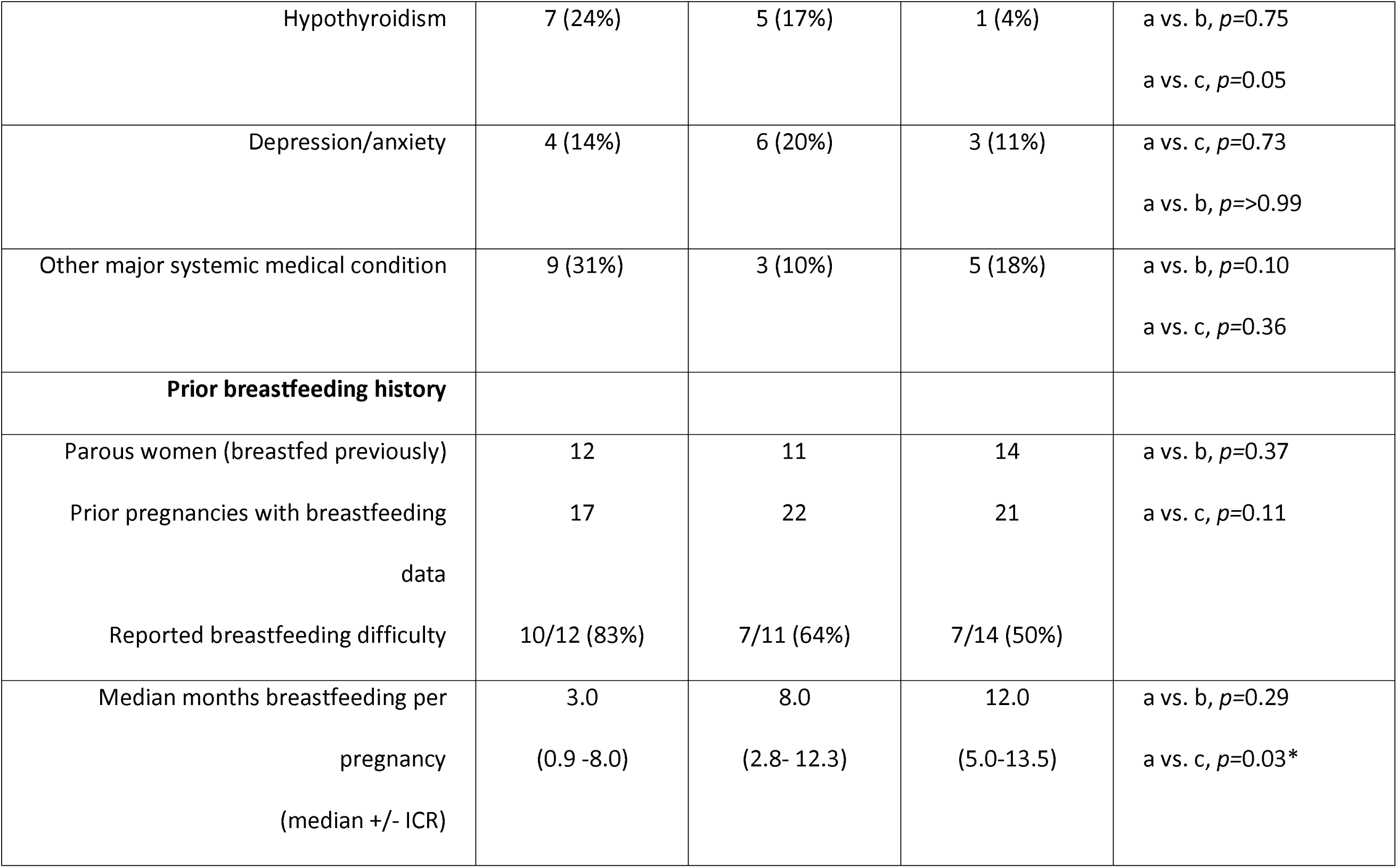
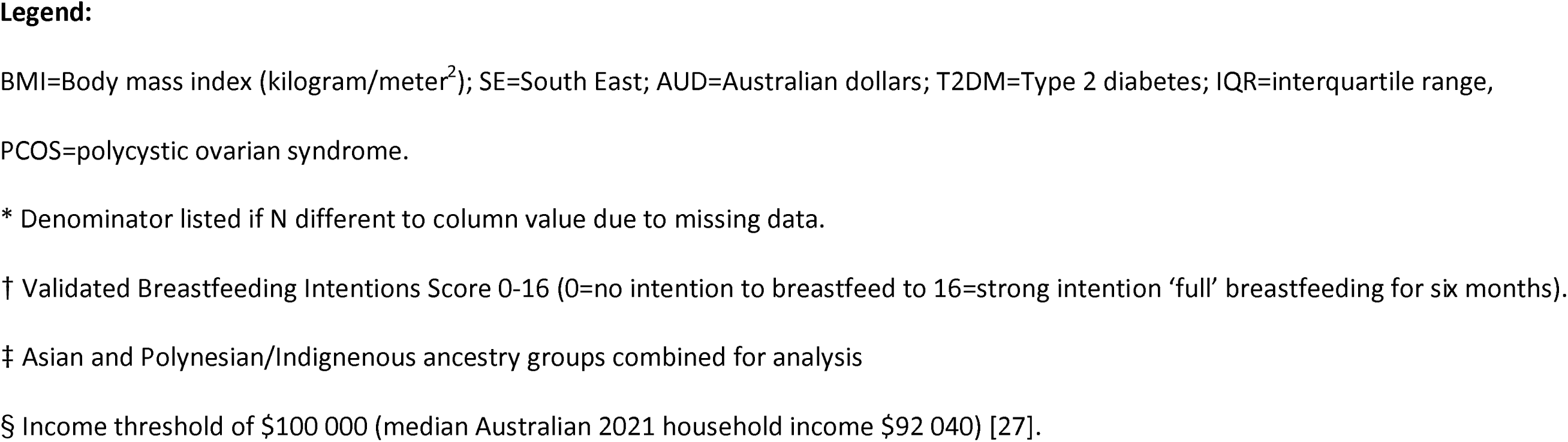
Baseline Data.

### 3.1 Baseline variables (Table 1)

There were no significant differences across many variables previously associated with breastfeeding establishment in the general population, including breastfeeding intention, education, parity and smoking. Unsurprisingly, given known ancestry differences in type 2 diabetes prevalences, there were more women of Asian ancestry in the type 2 diabetes group, compared with either of the two control groups. Women with type 2 diabetes were more likely to have an income below $AU100 000 compared to either control group.

### 3.2 Peripartum Maternal and Neonatal Outcomes (Table 2)

Women with type 2 diabetes gave birth more commonly by Caesarean section compared to BMI-matched controls. No significant difference was seen when compared to normal-BMI controls. Otherwise, there were no significant differences between obstetric variables including analgesia in labour, regional anaesthesia, and postpartum haemorrhage across the three groups. The mean postpartum hospital stay was significantly longer for women with type 2 diabetes than either control group.

**Table 2.**
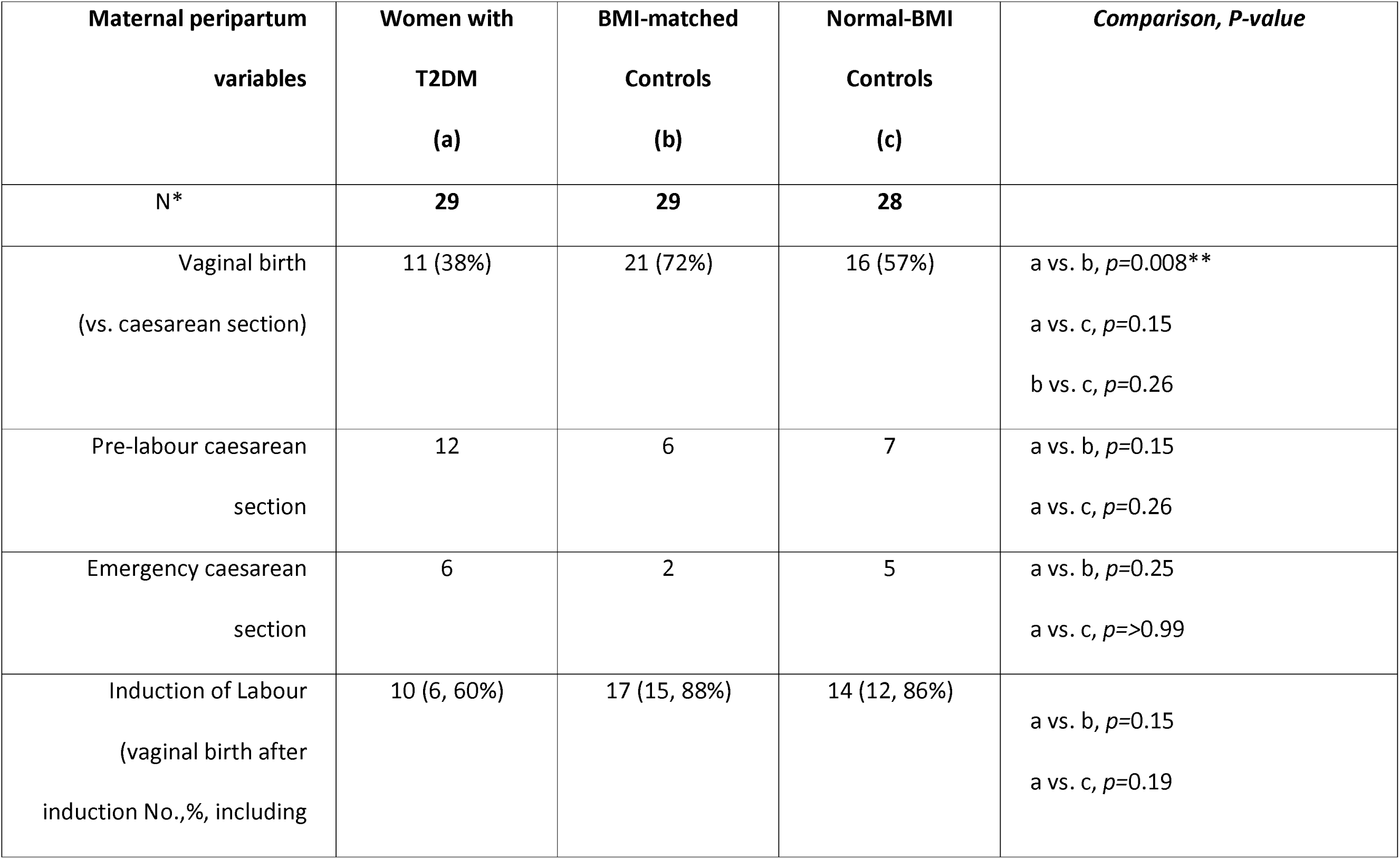
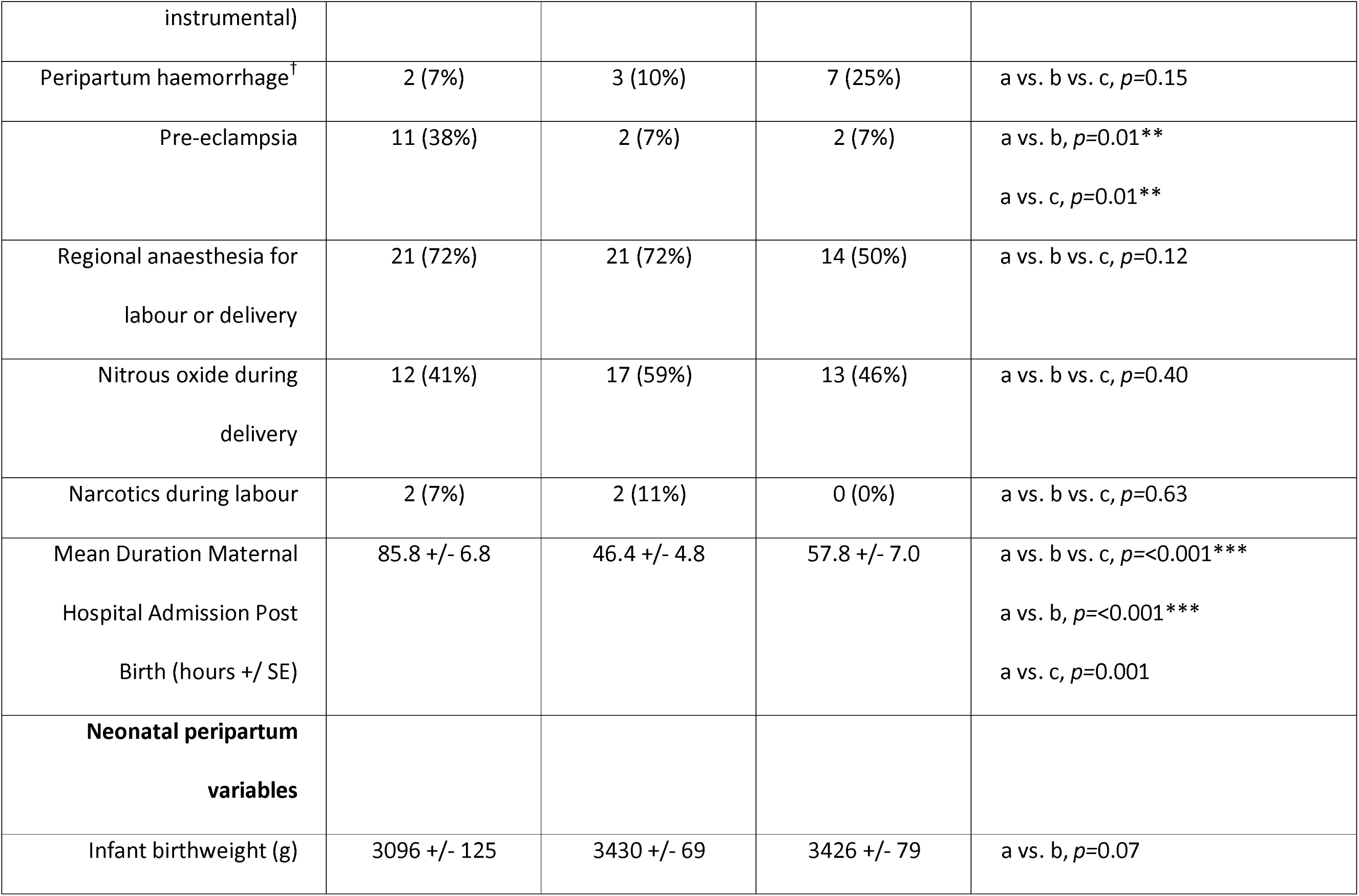
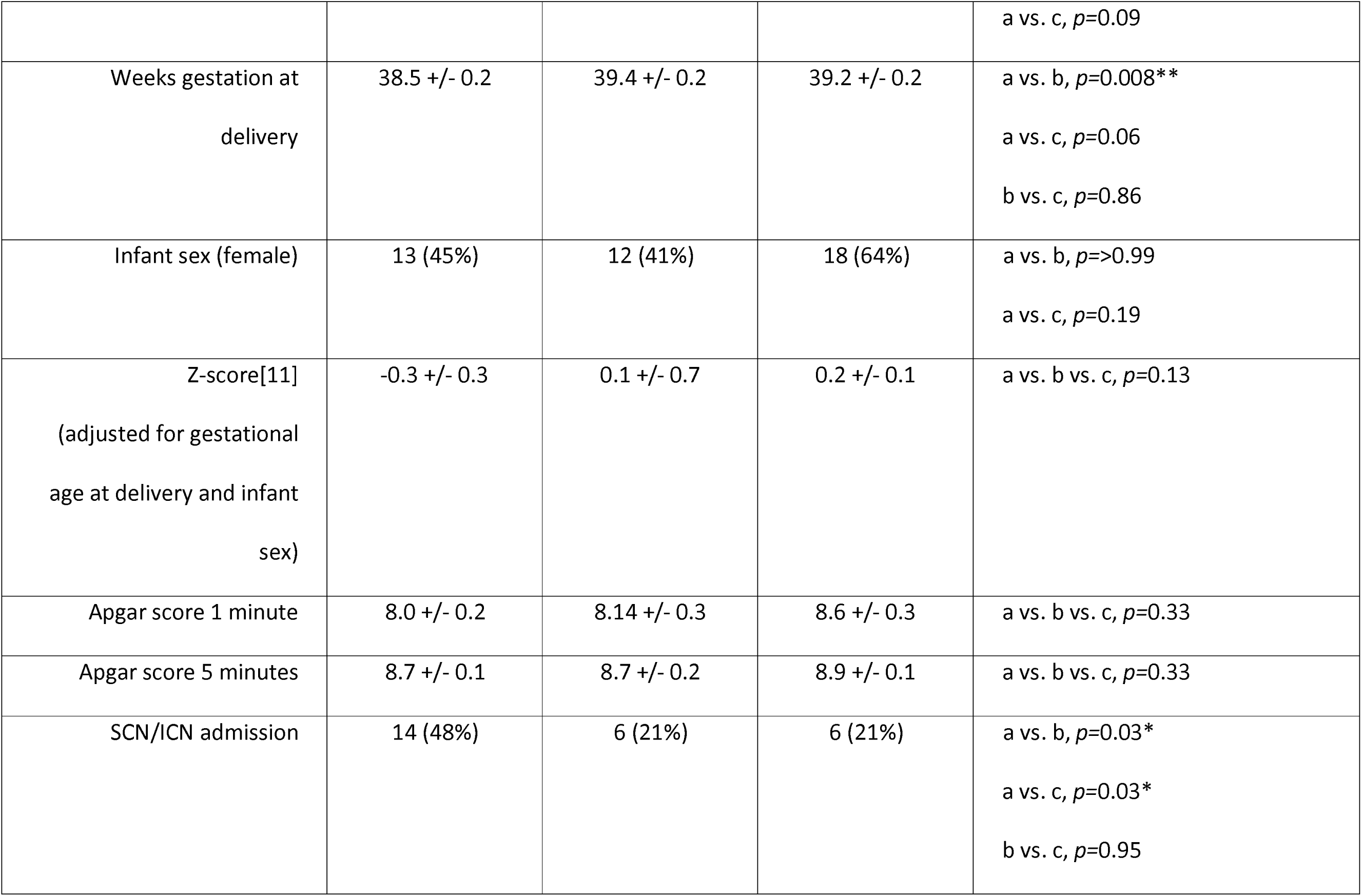
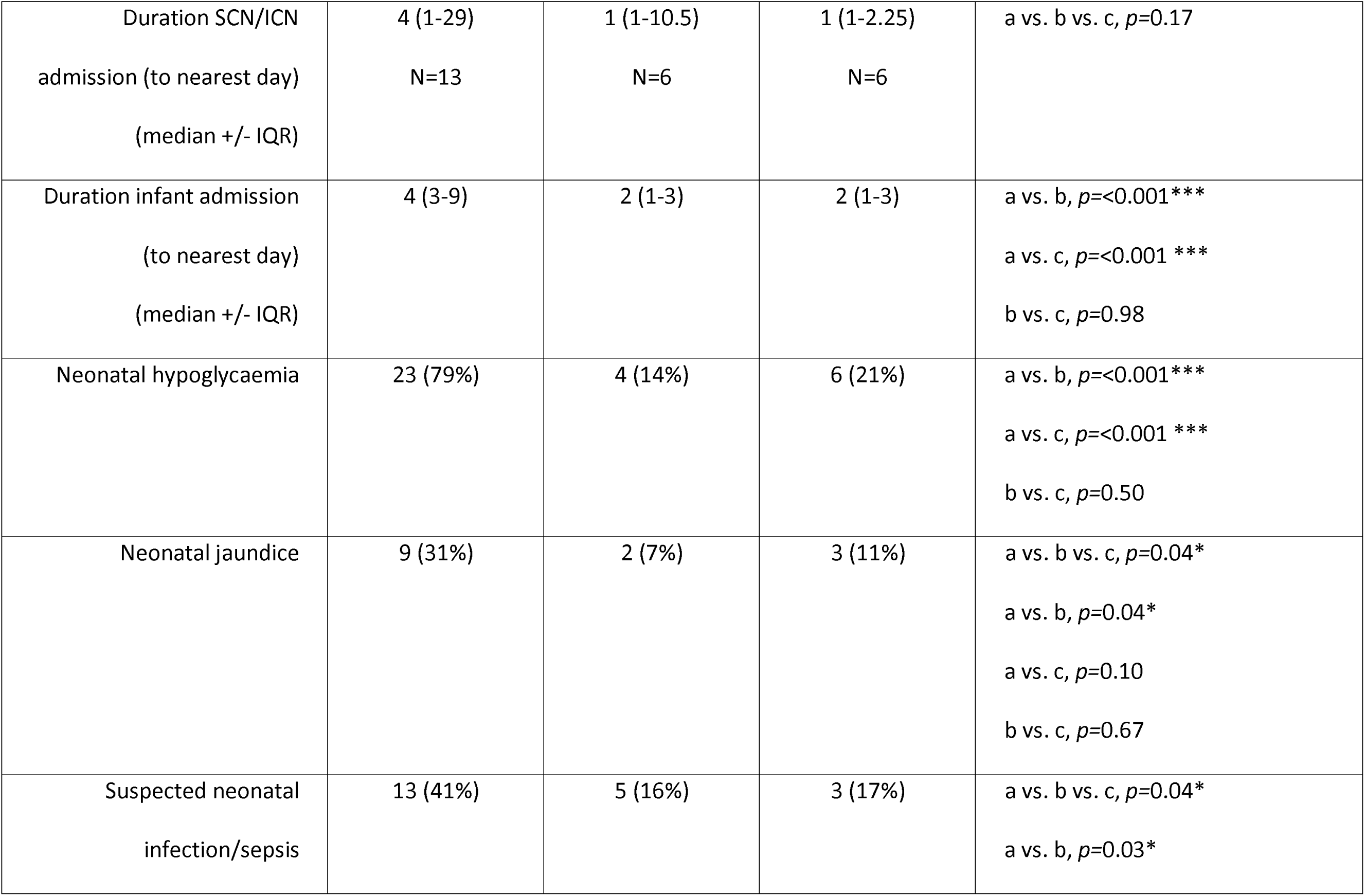
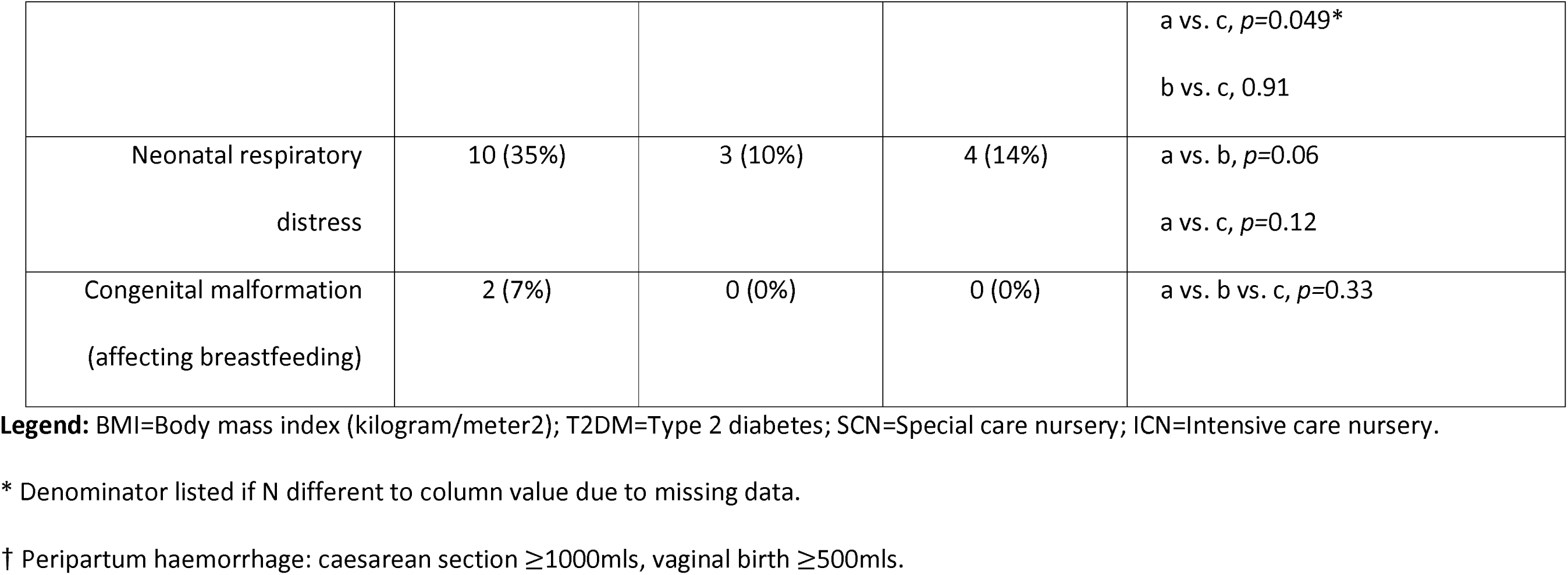
Peripartum Data.

| **Maternal peripartum**<br>**variables** | **Women with**<br>**T2DM**<br>**(a)** | **BMI-matched**<br>**Controls**<br>**(b)** | **Normal-BMI**<br>**Controls**<br>**(c)** | ***Comparison, P-value*** |
| --- | --- | --- | --- | --- |
| **N\*** | **29** | **29** | **28** |  |
| Vaginal birth<br>(vs. caesarean section) | 11 (38%) | 21 (72%) | 16 (57%) | a vs. b, $p=0.008^{**}$<br>a vs. c, $p=0.15$<br>b vs. c, $p=0.26$ |
| Pre-labour caesarean<br>section | 12 | 6 | 7 | a vs. b, $p=0.15$<br>a vs. c, $p=0.26$ |
| Emergency caesarean<br>section | 6 | 2 | 5 | a vs. b, $p=0.25$<br>a vs. c, $p=>0.99$ |
| Induction of Labour<br>(vaginal birth after<br>induction No.,%, including | 10 (6, 60%) | 17 (15, 88%) | 14 (12, 86%) | a vs. b, $p=0.15$<br>a vs. c, $p=0.19$ |
| instrumental) |  |  |  |  |
| Peripartum haemorrhage<sup>†</sup> | 2 (7%) | 3 (10%) | 7 (25%) | a vs. b vs. c, $p=0.15$ |
| Pre-eclampsia | 11 (38%) | 2 (7%) | 2 (7%) | a vs. b, $p=0.01^{**}$<br>a vs. c, $p=0.01^{**}$ |
| Regional anaesthesia for<br>labour or delivery | 21 (72%) | 21 (72%) | 14 (50%) | a vs. b vs. c, $p=0.12$ |
| Nitrous oxide during<br>delivery | 12 (41%) | 17 (59%) | 13 (46%) | a vs. b vs. c, $p=0.40$ |
| Narcotics during labour | 2 (7%) | 2 (11%) | 0 (0%) | a vs. b vs. c, $p=0.63$ |
| Mean Duration Maternal<br>Hospital Admission Post<br>Birth (hours +/- SE) | 85.8 +/- 6.8 | 46.4 +/- 4.8 | 57.8 +/- 7.0 | a vs. b vs. c, $p<0.001^{***}$<br>a vs. b, $p<0.001^{***}$<br>a vs. c, $p=0.001$ |
| **Neonatal peripartum**<br>**variables** |  |  |  |  |
| Infant birthweight (g) | 3096 +/- 125 | 3430 +/- 69 | 3426 +/- 79 | a vs. b, $p=0.07$ |
| | | | | a vs. c, $p=0.09$ |
| Weeks gestation at delivery | 38.5 +/- 0.2 | 39.4 +/- 0.2 | 39.2 +/- 0.2 | a vs. b, $p=0.008^{**}$<br>a vs. c, $p=0.06$<br>b vs. c, $p=0.86$ |
| Infant sex (female) | 13 (45%) | 12 (41%) | 18 (64%) | a vs. b, $p=>0.99$<br>a vs. c, $p=0.19$ |
| Z-score[11]<br>(adjusted for gestational age at delivery and infant sex) | -0.3 +/- 0.3 | 0.1 +/- 0.7 | 0.2 +/- 0.1 | a vs. b vs. c, $p=0.13$ |
| Apgar score 1 minute | 8.0 +/- 0.2 | 8.14 +/- 0.3 | 8.6 +/- 0.3 | a vs. b vs. c, $p=0.33$ |
| Apgar score 5 minutes | 8.7 +/- 0.1 | 8.7 +/- 0.2 | 8.9 +/- 0.1 | a vs. b vs. c, $p=0.33$ |
| SCN/ICN admission | 14 (48%) | 6 (21%) | 6 (21%) | a vs. b, $p=0.03^{*}$<br>a vs. c, $p=0.03^{*}$<br>b vs. c, $p=0.95$ |
| Duration SCN/ICN admission (to nearest day)<br>(median +/- IQR) | 4 (1-29)<br>N=13 | 1 (1-10.5)<br>N=6 | 1 (1-2.25)<br>N=6 | a vs. b vs. c, $p=0.17$ |
| Duration infant admission (to nearest day)<br>(median +/- IQR) | 4 (3-9) | 2 (1-3) | 2 (1-3) | a vs. b, $p<0.001^{***}$<br>a vs. c, $p<0.001^{***}$<br>b vs. c, $p=0.98$ |
| Neonatal hypoglycaemia | 23 (79%) | 4 (14%) | 6 (21%) | a vs. b, $p<0.001^{***}$<br>a vs. c, $p<0.001^{***}$<br>b vs. c, $p=0.50$ |
| Neonatal jaundice | 9 (31%) | 2 (7%) | 3 (11%) | a vs. b vs. c, $p=0.04^*$<br>a vs. b, $p=0.04^*$<br>a vs. c, $p=0.10$<br>b vs. c, $p=0.67$ |
| Suspected neonatal infection/sepsis | 13 (41%) | 5 (16%) | 3 (17%) | a vs. b vs. c, $p=0.04^*$<br>a vs. b, $p=0.03^*$ |
| | | | | a vs. c, $p=0.049^*$<br>b vs. c, 0.91 |
| Neonatal respiratory<br>distress | 10 (35%) | 3 (10%) | 4 (14%) | a vs. b, $p=0.06$<br>a vs. c, $p=0.12$ |
| Congenital malformation<br>(affecting breastfeeding) | 2 (7%) | 0 (0%) | 0 (0%) | a vs. b vs. c, $p=0.33$ |

**Legend:** BMI=Body mass index (kilogram/meter<sup>2</sup>); T2DM=Type 2 diabetes; SCN=Special care nursery; ICN=Intensive care nursery.
\* Denominator listed if N different to column value due to missing data.
† Peripartum haemorrhage: caesarean section $\geq 1000$ mls, vaginal birth $\geq 500$ mls.

Infants of mothers with type 2 diabetes were delivered at an earlier gestation than both control groups, although the z score (adjusted measure of neonatal weight for gestation and sex)[11] did not differ significantly between groups. These infants were also more likely to have jaundice, infection/sepsis, admittance to the special or intensive care nursery, treatment for hypoglycaemia, and a longer hospital stay compared with infants of mothers in either control group.

### 3.3 Feed Type and Frequency During Hospital Admission (See **Table 3**)

Women with type 2 diabetes breastfed fewer times in the first 24 hours postpartum (4.4+/-0.8 times in 24 hours versus 7.8+/-0.6, P=0.009 and 8.5+/-0.6, P=0.001 times in 24 hours) in the BMI-matched and normal-BMI groups respectively. In addition, 80% of their infants received formula feeds in hospital compared to 24% of the BMI-matched group (P=<0.001) and 29% of the normal-BMI group (P=<0.001).

**Table 3.** Postpartum Breastfeeding Behaviour.

| **Infant feeding variables** | **Women with**<br>**T2DM**<br>**(a)** | **BMI-matched**<br>**Controls**<br>**(b)** | **Normal-BMI**<br>**Controls**<br>**(c)** | **Comparison, P-value** |
| --- | --- | --- | --- | --- |
| Breastfeeding 'full'<sup>†</sup> at<br>discharge | 8 (28%) | 19 (68%) | 25 (89%) | a vs. b, $p=0.003^{**}$<br>a vs. c, $p=<0.001^{***}$<br>b vs. c, $p=0.10$ |
| Breastfeeding 'full' four<br>months | 7 (24%) | 11 (38%) | 18 (64%) | a vs. b, $p=0.26$<br>a vs. c, $p=0.002^{**}$<br>b vs. c, $p=0.049^{*}$ |
| Breastfeeding<br>'full/mostly'<sup>‡</sup> at four<br>months | 11 (38%) | 13 (45%) | 20 (71%) | a vs. b, $p=0.59$<br>a vs. c, $p=0.01^{*}$<br>b vs. c, $p=0.04^{*}$ |
| Breastfeeding 'any'<sup>§</sup> at<br>four months | 17 (63%) | 15 (54%) | 23 (82%) | a vs. b, $p=0.60$<br>a vs. c, $p=0.049^{*}$ |
| | | | | b vs. c, $p=0.04^*$ |
| Breastfed within 1 hour of delivery | 21 (72%) | 27 (93%) | 26 (93%) | a vs. b, $p=0.08$<br>a vs. c, $p=0.08$ |
| Mean breastfeeding episodes first 24 hours<br>(women admitted $\geq 24$ hours) | 4.4 +/- 0.8<br>N=27 | 7.8 +/-0.6<br>N=25 | 8.5 +/- 0.6<br>N=24 | a vs. b, $p=0.009^{**}$<br>a vs. c, $p=0.001^{**}$ |
| Mean number of breastfeeds/expressed breastfeeds first 24 hours | 5.3 +/- 0.9 | 9.6 +/-0.6 | 9.0 +/- 0.7 | a vs. b, $p=<0.001^{***}$<br>a vs. c, $p=0.003^*$ |
| Number of breastfeeds in infants able to breastfeed (BF >0) and admitted first 24 hours | 7.1+/-0.7<br>N=17 | 8.5+/- .5<br>N=23 | 9.3 +/- 0.4<br>N=22 | a vs. b, $p=0.25$<br>a vs. c, $p=0.03^*$ |
| Neonates per group supplemented formula | 23 (80%) | 7 (24%) | 8 (29%) | a vs. b, $p=<0.001$<br>a vs. c, $p=<0.001$ |
| during admission |  |  |  |  |
| Supplemented with<br>formula first 24 hours<br>(excluded if discharged before<br>24 hours) | 13 (48%)<br>N=27 | 4 (17%)<br>N=24 | 5 (21%)<br>N=24 | a vs. b, $p=0.04$<br>a vs. c, $p=0.09$ |
| Mean formula feeds first<br>24 hours (if formula given) | 3.5+/-0.4<br>N=12 | 3.5+/-1.5<br>N=4 | 4.0+/-1.3<br>N=5 | a vs. b vs. c, $p=0.90$ |

**Legend:** Peripartum data are shown for all women with data collected at all three time points. BMI=Body mass index (kilogram/meter<sup>2</sup>);
T2DM=Type 2 diabetes.
\* Denominator listed if N different to column value due to missing data.
† 'full' refers to only breastmilk given without other liquids or solids except for vitamins, minerals or medicines at the time of survey.
‡ 'mostly' refers to a greater than 50% of infant's requirements are provided via breastmilk.
§ 'any' refers to any combination of breastmilk and formula, including 'full' breastfeeding.

Main reasons for formula initiation are shown in Table 4. For the 23 (of 29) women with type 2 diabetes who used formula in hospital, the most common reason for first use was neonatal hypoglycaemia management in 61% (14 of the 23 infants fed formula). Hypoglycaemia was a less common reason for first formula use in the BMI-matched group (3 of 7 [43%] infants, P=0.003) and normal-BMI group (1 of 8 [13%] infants, P= <0.001).

**Table 4.**
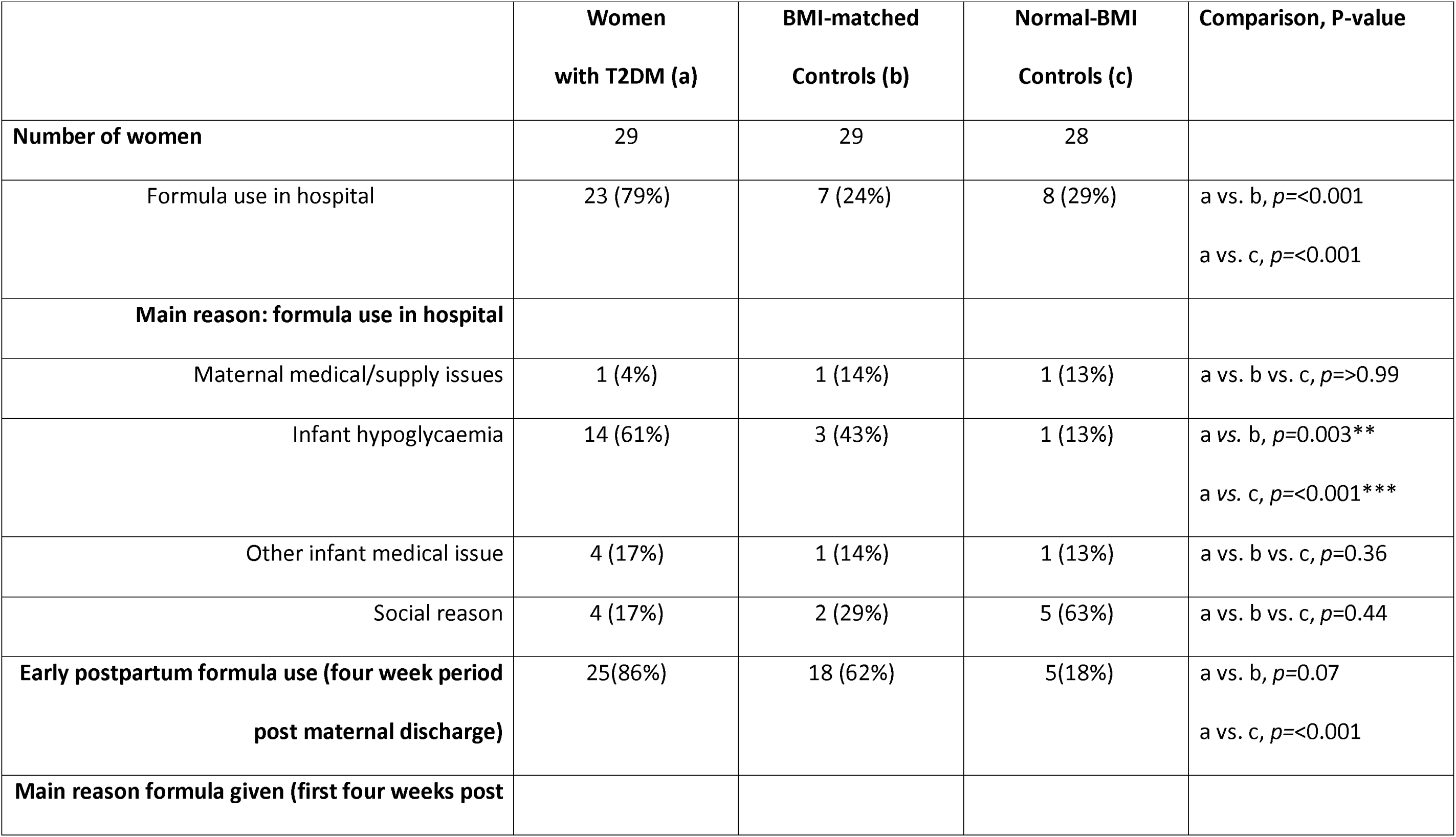
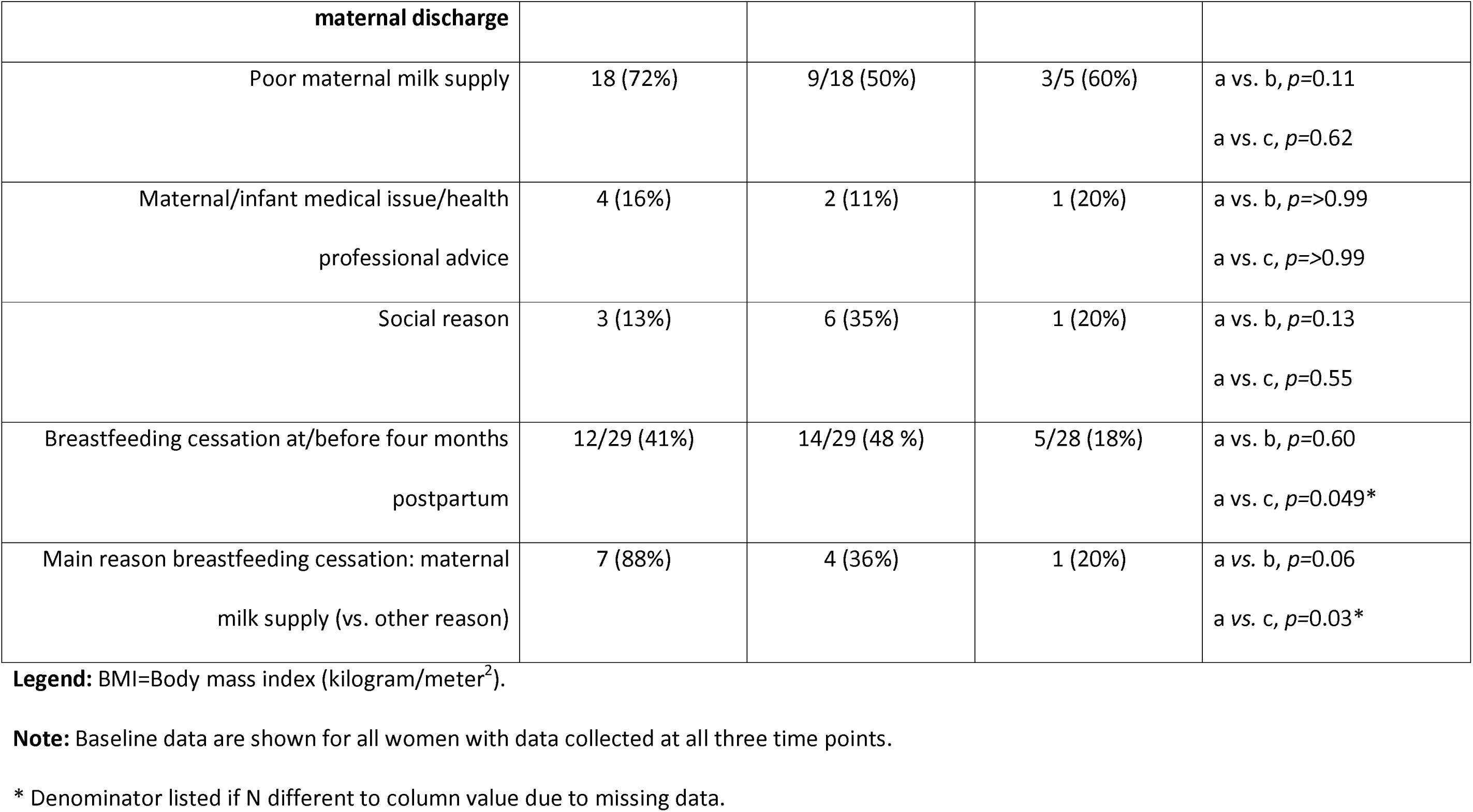
Reasons For Formula Use and/or Breastfeeding Cessation.

### 3.4 Breastfeeding At Hospital Discharge and Four Months Postpartum (Table 3)

Women with type 2 diabetes were less likely to be ‘full’ breastfeeding at discharge than BMI-matched or normal-BMI groups (8 of 29 [28%] vs 19 of 29 68%, P=<0.001 and 25 of 28 [89%], P=<0.001) respectively.

At four months postpartum, women with type 2 diabetes and BMI-matched controls were less likely to report ‘full’ or ‘any’ breastfeeding than normal-BMI controls. However, ‘full’ and ‘any’ breastfeeding rates were similar between women with type 2 diabetes and BMI-matched controls. Findings were also similar when comparing women ‘full’ or ‘mostly’ breastfeeding versus more than 50% formula feeding (terms used to differentiate between occasional formula use for convenience [‘mostly’ breastfeeding] versus inadequate milk supply [feeding at least 50% formula]). By four months postpartum, 12 of 29 (41%) women with type 2 diabetes, 14 of 29 (48%) BMI-matched controls, and 5 of 28 (18%) of normal BMI women had ceased breastfeeding altogether.

### 3.5 Reasons for Formula Use Post Hospital Discharge and Breastfeeding Cessation Prior to Four Months Postpartum (Table 4)

Primary reasons for formula use in the first four weeks after maternal discharge and for breastfeeding cessation prior to four months are outlined in Table 4. The most common reason given for formula use in the first four weeks post hospital discharge in all three groups was perceived poor supply of breastmilk. This was a significantly more common reason for breastfeeding cessation prior to four months in the type 2 diabetes group compared to the normal-BMI group (88% vs. 20%, P=0.03), with a trend to difference noted between the type 2 diabetes and normal-BMI groups (36% vs. 14%, P=0.06).

### 3.6 Variables Affecting Breastfeeding Four Months Postpartum in Women with Type 2 Diabetes (Table S1 Supplementary Data)

No demographic differences were observed between women with type 2 diabetes who were and were not ‘full’ breastfeeding at four months, noting the small ‘full’ breastfeeding group and limited sample size. Women who were ‘full’ breastfeeding at four months were more likely to have been ‘full’ breastfeeding at discharge. None had an infant with a prolonged neonatal nursery admission or used formula in the first 24 hours. ‘Full’ breastfeeding at four months was not associated with antenatal metformin use (Table S1) or insulin dose/kg at delivery quantified by linear regression (OR 1.3, CI 0.6–3.0; P=0.44)(data not shown).

## 4.0 Discussion

This is the first prospective study we are aware of that compares long-term breastfeeding in women with type 2 diabetes to matched controls. We have demonstrated that ‘full’ and ‘any’ breastfeeding rates four months postpartum are similar in women with type 2 diabetes and normoglycaemic BMI-matched controls (for ‘full’ breastfeeding: 24% vs. 38% [p=0.26]; for any: 63% vs. 54% [P=0.6]). In contrast, rates of breastfeeding were significantly higher (approximately 2.7-fold and 1.3-fold respectively) for ‘full’ or ‘any’ breastfeeding in normal BMI controls, compared to women with type 2 diabetes. Similarly, rates of ‘full’ or ‘any’ breastfeeding were again significantly higher (1.7-fold, and 1.5-fold, respectively) in normal BMI controls compared with BMI-matched controls. This suggests type 2 diabetes may not confer any additional risk of early breastfeeding cessation beyond that seen from elevated BMI.

Only two prior studies have assessed the independent effects of BMI and type 2 diabetes on breastfeeding: a retrospective cohort[4] and a prospective rural Australian study[5]. Both found elevated BMI, but not type 2 diabetes, independently predicted reduced ongoing breastfeeding, consistent with our results. The Pregnancy and Neonatal Diabetes Outcomes in Remote Australia (PANDORA) study is the only prospective comparison of long-term breastfeeding in type 2 diabetes versus normoglycaemic controls. At six months, predominant breastfeeding occurred in 58% of Indigenous women with type 2 diabetes vs 77% of controls, and 40% of non-Indigenous women with type 2 diabetes vs 60% of controls[5]. Differences were not significant. This contrasts with our findings, possibly due to different timepoints and a rural, predominantly Indigenous cohort versus our urban population.

In-hospital formula use increases early breastfeeding cessation in women intending to exclusively breastfeed[12]. However, many studies do not differentiate between occasional formula use (e.g., to allow a time away from baby without pumping) from regular formula use to manage inadequate supply; and most studies define breastfeeding in both scenarios as ‘combined formula/breastmilk’ feeding. Our study allowed for this by differentiating women who were ‘full or mostly’’ breastfeeding from those women feeding at least 50% formula. Nevertheless, our study still demonstrated lower rates of ‘full or mostly’ breastfeeding in the women with type 2 diabetes and BMI-matched controls compared to normal-BMI matched controls.

Although numbers of women ‘full’ breastfeeding were not different in women with type 2 diabetes and the BMI-matched group, it is possible that relevant mediators were different. Type 2 diabetes and BMI-matched groups were similar across many parameters known to affect breastfeeding rates including education level, smoking and income[13]; but type 2 diabetes women delivered an earlier gestation, more commonly by caesarean; and their infants were more likely to have hypoglycaemia and receive more formula and fewer breastfeeds in the first 24 hours postpartum than the BMI-matched group. Earlier gestation at delivery and caesarean section were both independently associated with a reduced rate of breastfeeding at six months in a previous study examining breastfeeding in women with type 2 diabetes [5]. In-hospital formula use has also been shown to reduce full breastfeeding post-discharge by 50% in women without diabetes[14].

In addition, there was a trend to BMI-matched women to be more likely to start formula in the first four weeks postpartum for social reasons, and to be less likely to introduce formula for supply reasons, than women with type 2 diabetes. In contrast, there was a trend to women with type 2 diabetes being more likely to report poor supply as the primary reason for breastfeeding cessation than both BMI-matched and normal-BMI groups. Previous studies of breastfeeding in type 2 diabetes with normoglycaemic controls did not examine reasons for starting formula and/or ceasing breastfeeding[4, 5]. Although not formally assessed in this study, it is possible knowledge about the metabolic benefits of breastfeeding in diabetes mitigated breastfeeding cessation for social reasons in the type 2 diabetes group.

### 4.1 Role of Ancestry

More women in the type 2 diabetes group, compared with the control groups, were of South or Southwest Asian ancestry. Ancestry has been associated with differences in breastfeeding rates in the general community, with multiple possible contributions, including cultural values, community breastfeeding support, and/or differential acceptance of early formula use[5, 15, 16]. A recent Australian study found women from South and Southwest Asia introduce formula later and breastfeed for two months longer than their counterparts of European-Australian ancestry[17]. Given women of South and Southwest Asian ancestry in Australia are more likely to breastfeed, this suggests that our estimates for breastfeeding in type 2 diabetes might be influenced by the higher percentage of women of South and Southwest Asian ethnicity in this group, but is unlikely to explain the lower rate of breastfeeding seen in this group. The impact of ethnicity on breastfeeding in women with type 2 diabetes needs to be examined in larger studies.

### 4.2 Hormonal Factors

It has been postulated that insulin resistance may negatively affect breastmilk supply, and thereby contribute to poorer breastfeeding success, as reported previously in women with PCOS[18], obesity[19], gestational diabetes[20] and type 2 diabetes [21] and in obese bovine[22] and rodent models[23]. However, we did not observe a relationship with breastfeeding at four months postpartum and insulin dose immediately prepartum (a surrogate measure of insulin resistance) in women with type 2 diabetes. Further, antenatal metformin use was also not associated with ‘full’ breastfeeding at four months. Lastly, our BMI-matched controls all had negative oral glucose tolerance tests, so represent a relatively insulin-sensitive high BMI group; yet these women had similarly low breastfeeding rates to the type 2 diabetes group. Our data do not support a relationship between insulin resistance and breastfeeding.

### 4.3 Formula Feeding in Hospital and Full Breastfeeding at Discharge

In-hospital formula use and lack of ‘full’ breastfeeding at discharge predict reduced long-term ‘full’ breastfeeding in the general population[24]; whether this holds in type 2 diabetes is uncertain. In our cohort, women with type 2 diabetes had longer admissions, potentially increasing lactation support, yet formula use was common, often triggered by hypoglycaemia. Despite strong antenatal intent, only 29% were fully breastfeeding at discharge versus 72.4% nationally[25], and discharge status predicted continuation.

### 4.4 Strengths and Weakness

This is one of very few prospective studies examining breastfeeding rate in type 2 diabetes, with age- and parity-matched normoglycaemic controls, and the only study we are aware of that has a concurrent BMI-matched control. Unmatched parameters were also similar between groups. Further, GDM was excluded in our BMI-matched controls, allowing assessment of elevated BMI in women without overt insulin resistance. Our data collection was comprehensive and granular, including multiple parameters previously known to affect breastfeeding. In addition, long-term breastfeeding was measured, rather than them more common measure of breastfeeding at discharge. The degree of breastfeeding at four months postpartum was also quantified which is omitted from many studies.

However, overall our study size is small. There was loss to follow up in all three groups, precluding the pairwise statistical analysis that had initially been planned. Further, our study groups were not matched for ancestry. A limitation in our study was that only women with intention to breastfeed were recruited; thus our study illustrates the rate of ongoing breastfeeding in women with type 2 diabetes and matched controls intending to breastfeed, rather the rate of breastfeeding in all women with type 2 diabetes. This may give an over-estimate of breastfeeding rates in all three groups, although as nearly all (96%) of women in Australia intend to initiate breastfeeding[26] this limitation may have minimal impact on our findings.

### 4.5 Future Directions

Larger studies controlling for income and ancestry are important to clarify the impact of these variables, as well as assessing the relationships between postpartum variables such as formula feeding, number of early breastfeeding episodes, neonatal illness and nursery admission. Reasons for breastfeeding cessation also requires greater exploration. The strong association of formula use in hospital and ‘full’ breastfeeding at discharge on long-term breastfeeding continuation suggests an urgent need to investigate ways to improve ‘full’ breastfeeding at discharge in women with type 2 diabetes. Our findings suggest ‘full’ breastfeeding at discharge may be a useful proxy for continued breastfeeding in women with type 2 diabetes in future studies.

### 4.6 Conclusion

In summary, our study showed that both women with type 2 diabetes and BMI-matched women had reduced rates of ‘full’ and ‘any’ breastfeeding at four months postpartum compared to normal-BMI women. Important peripartum factors associated with full breastfeeding at four months included less formula use in hospital, early breastfeeding episodes and ‘full’ breastfeeding at discharge. In women with type 2 diabetes, the rate of ‘full’ breastfeeding at four months was only 24%, despite high maternal breastfeeding intentions. Further research is urgently needed to identify strategies for improving in hospital breastfeeding in women with type 2 diabetes.

## Data Availability

Data is available from the authors on reasonable request.

## Acknowledgments

The authors would like to thank all the study participants.

## Author Contributions

Conceptualisation FLB, ELD and LKC.; Statistical analysis FLB. Roles/Writing - original draft FB, ELD and LKC; and Writing - review & editing FLB, ELD and LCK. article.

## Declaration of Generative AI and AI-assisted technologies in the Manuscript Preparation Process

During the preparation of this work the author(s) used ChatGPT 5.2 to reduce word count. After using this tool/service, the author(s) reviewed and edited the content as needed and take(s) full responsibility for the content of the published article.

