## Supplementary Files for "A Prospective Controlled Study of Postpartum Breastfeeding Rates in Women with Type 2 Diabetes: Does Obesity Matter More Than Diabetes?"

Supplementary Files: Table S1 and Survey Tools S2, S3 and S4

**Table S1 -** **Variables Associated with Full Breastfeeding at Four Months Postpartum in Women with Type 2 Diabetes**

| **Maternal variables** | **Full BF**  **(N=7)*** | **Not Full BF**  **(N=22)** | ***Comparison, P-value*** |
| --- | --- | --- | --- |
| Age at due date (≥ 35 years) | 3 (43%) | 8 (36%) | >0.99 |
| Parity (multiparous) | 4 (57%) | 12 (55%) | >0.99 |
| BMI ≥ 30 | 4 (57%) | 15 (68%) | 0.66 |
| Maternal education (Degree or higher) | 3 (43%) | 12 (55%) | 0.68 |
| Household income >100 000K^†^ | 6 (86%) | 15 (68%) | 0.63 |
| Maternal ancestry  Asian  European | 2 (29%)  5 (71%) | 11 (50%)  11 (50%) | 0.41 |
| South or Southwest Asian ancestry | 2 (29%) | 5 (23%) | 0.68 |
| **Infant variables** | | | |
| Breastfed within one hour postpartum | 5 (71%) | 11 (69%) | 0.41 |
| No breastfeeding first 24 hours postpartum | 3 (43%) | 16 (73%) | 0.19 |
| Number breastfeeds first 24 hours > or equal to 8 | 3/6 (50%) | 3 (14%) | 0.35 |
| Full Breastfeeding at Discharge | 6 (86%) | 2 (9%) | <0.001*** |
| Formula in the first 24 hours | 0 (0%) | 13 (59%) | 0.008** |
| Formula during neonatal admission | 3 (43%) | 21 (95%) | 0.007** |
| Neonatal hypoglycaemia | 4 (57%) | 19 (86%) | 0.08 |
| Neonatal nursery admission (N= 14) | 4 (57%) | 10 (45%) | 0.68 |
| Neonatal nursery admission > 3 days (N=7) | 0 (0%) | 7 (32%) | 0.02* |
| Infant admission >3 days (N=18) | 4 (57%) | 14 (64%) | >0.99 |
| Neonatal jaundice | 1 (14%) | 8 (36%) | 0.38 |
| Neonatal sepsis or infection | 3 (43%) | 10 (45%) | >0.99 |
| Composite neonatal infection, jaundice, hypoglycemia | 4 (57%) | 21 (95%) | 0.03* |
| Neonatal respiratory Distress | 2 (29%) | 8 (36%) | >0.99 |
| Metformin use during pregnancy | 3 (43%) | 16 (73%) | 0.19 |

**Legend:** BMI=Body mass index (kilogram/meter^2)^; T2DM=Type 2 diabetes; SCN=Special care nursery; ICN=Intensive care nursery; Groups: a=T2DM, b=BMI-matched, c=normal-BMI.

*Denominator listed if N different to column value due to missing data

† Income threshold of $100 000 (median Australian 2021 household income $92 040).

**(S2) Breastfeeding in Mothers with Type 2 Diabetes: Initial Survey**

(Clinical survey to be administered by researcher)

Current date	/	/	(day/month/year)

Current gestation/Expected date confinement

Patient Code

1. What is your date of birth?	___/___/____

day	month	year

2. Are you currently……

☐_1_ Married

☐_2_ Living with partner/defacto

☐_3_ Single, not living with a partner

☐_4_ Single, living with my family

3. What is the highest qualification you have completed?

☐_1_ Less than Year 10

☐_2_ Year 10 or equivalent (e.g. School Certificate)

☐_3_ Year 12 or equivalent (e.g. Higher School Certificate)

☐_4_ Trade/apprenticeship (e.g. hairdresser, chef)

☐_5_ Certificate/diploma (e.g. childcare, technician, laboratory)

☐_6_ University degree

☐_7_ Higher university degree (e.g. Graduate Diploma, Masters, PhD)

4. Which of the following best describes your main current employment status?

☐_1_ In full-time paid work

☐_2_ In part-time or casual paid work

☐_3_ Not in paid employment

5. What was the average total income (before tax) of your household from all sources in the last financial year? (eg. You and your partner) Include pensions and allowances.

☐_0_ Nil/Negative income

☐_1_ $0 - $385 per week ($0 - $20 000 per year)

☐_2_ $386 - $673 per week ($20 001 - $35 000 per year)

☐_3_ $674 - $961 per week ($35 001 - $50 000 per year)

☐_4_ $962 - $1346 per week ($50 001 - $70 000 per year)

☐_5_ $1347 - $1923 per week ($70 0001 - $100 000 per year)

☐_6_ more than $1923 per week (more than $100 000 per year)

☐_7_ Not sure	☐_8_ Prefer not to answer

1. Does your family have a Health Care Card? (this is different from a Medicare or Private	Health Insurance card)	No ☐_1_ Yes ☐_2_
2. Which country were you born in?

☐ Australia ☐ Another country (please specify)

3. Do you speak a language other than English at home?

No ☐_1_ Yes ☐_2_ (please specify)

4. Are you of Aboriginal or Torres Strait Islander origin? No ☐_1_ Yes ☐_2_
5. Do you have children living in your household? No ☐_1_ Yes ☐_2_ (how many)
6. What is the post-code in the suburb you live in?
7. What was your pre-pregnancy weight in kilograms?
8. What is your height in centimeters?	Calculated BMI=
9. How many pregnancies and children have you had? (Parity described as GPMT)

| G | P | T | M |
| --- | --- | --- | --- |

10. Do you have any medical problems? Please give further details

|  | Condition |
| --- | --- |
| Heart/Cardiovascular |  |
| Lungs |  |
| Digestive system |  |
| Kidneys |  |
| Brain/nerves |  |
| Skin |  |
| Bones/joints |  |
| Hormonal |  |
| Blood |  |
| Other |  |

1. Are you on any other medications? Please give details.

| Medication | Dose |
| --- | --- |

2. Feeding your baby. Please choose the answer that most closely matches your opinion, considering both your feeding plans and the likelihood you will carry out those plans.

|  | **(1)**  **Very much disagree** | **(2)**  **Some-what disagree** | **(3)**  **Unsure** | **(4)**  **Some-**  **what agree** | **(5)**  **Very much agree** |
| --- | --- | --- | --- | --- | --- |
| I am planning to only formula feed my baby (I will not breastfeed at all) |  |  |  |  |  |
| I am planning to at least give breastfeeding a try |  |  |  |  |  |
| When my baby is 1 month old, I will be breastfeeding without using any formula or other milk |  |  |  |  |  |
| When my baby is 3 months old, I will be breastfeeding without using any formula or other milk |  |  |  |  |  |
| When my baby is 6 months old, I will be breastfeeding without using any formula or other milk |  |  |  |  |  |

1. Have you breastfed previously?

☐_1_ Yes

☐_2_ No

2. Did you have difficulty initiating or continuing breastfeeding with prior children?

☐_1_ Yes

☐_2_ No

☐_3_ Not applicable (if never breastfed)

3. Do you remember when you ceased breastfeeding with prior children? Please give details.

| Child 1 |
| --- |
| Child 2 |
| Child 3 |
| Child 4 |

**(S3) Breastfeeding in Mothers with Type 2 Diabetes: Peripartum Data Collection**

Patient Code

Date of Delivery	dd/mm/yy

| Medical Conditions in pregnancy |
| --- |
| HBA1cs in pregnancy  -dates  -value |
| Weeks gestation at birth |
| Baby Weight at Birth |
| Mother’s Weight on admission prior to Birth |
| Type of Birth  -Induction  -SVD  -Assisted VD  -elective LSCS  -emergency LSCS  -time in labour prior to emLSCS |
| Maternal analgesia during labour/birth  -epidural  -pethidine  -nitrous oxide  -other (describe) |
| Maternal complications  -post partum haemorrhage  - perineal tear  - pre-eclampsia  -shoulder dystocia  -other (describe) |
| Fetal complications  -hypoglycaemia  -respiratory distress  - malformations  - hyperbilirubinaemia  -other (describe) |
| First breastfeed within 1 hour (yes/no)  Time post delivery if recorded |
| Skin to skin contact post-partum (yes/no)  Duration (minutes) |
| Mother baby separation during admission (yes/no)  Time post-partum and duration (minutes) |
| Were supplemental feeds given in hospital? (yes/no) |
| Were supplemental feeds with  -formula  -breastmilk  -formula and breastmilk  -Other (ie dextrose - describe) |
| -Time (post birth) first supplemental feed  -type of first supplemental feed |
| How many supplemental feeds given during hospital stay |
| Method of supplemental feed  -NG tube  -bottle  -Other (describe) |
| Length of stay in hospital (days) |
| Breastfeeding method at discharge  Breastfeeding= BF,  Mixed feeding = MF  Formula feeding= FF |

**(S4) Breastfeeding in Mothers with Type 2 Diabetes: Telephone Survey at 4 Months Post-partum**

Patient code

Current date	/	/	(day/month/year)

1. How old is your baby now? (weeks)
2. Prior to giving birth did you express colostrum for your baby?

No ☐_1_ Yes ☐_2_

3. Prior to giving birth did you receive education on expressing colostrum for your baby?

No ☐_1_ Yes ☐_2_

4. Did you ever breastfeed your baby either in hospital or after you went home?

No ☐_1_ Yes ☐_2_

5. When you left hospital how were you feeding your baby?

☐_1_ Breastfeeding only

☐_2_ Formula feeding only

☐_3_ Both breast and formula feeding

6. Are you currently

☐_1_ Breastfeeding only

☐_2_ Formula feeding only

☐_3_ Both breast and formula feeding

7. If you are both breast and formula feeding are you

☐_1_ Mostly breastfeeding

☐_2_ Mostly formula feeding

☐_3_ Around half breastfeeding and half formula feeding

8. Are you currently feeding your baby with any foods other than breastmilk or formula?

No ☐_1_ Yes ☐_2_

9. If yes, when did you start these solid foods? (weeks)
10. Did you take Motilium (domperidone) to increase your supply?

No ☐_1_ Yes ☐_2_

11. How long did you take it for (months)
12. Are you still taking it?

No ☐_1_ Yes ☐_2_

13. Did you take any other herbs or supplements to increase your milk supply? (please list)
14. Are you taking any other medications or supplements ? (Please list with doses)

15. If you are gave formula in the first 4 weeks after your baby was born what were the reasons for supplementing with formula?

(Indicate as many as apply and note if during admission (A) or post discharge (D).

| Baby had low blood sugars |
| --- |
| Baby was admitted to the special care or intensive care unit |
| Baby did not gain enough weight (or lost weight) |
| Baby was sick and could not breastfeed |
| Baby was premature and could not breastfeed |
| You thought you did not have enough milk supply |
| A health professional said you should for medical reasons |
| You were sick or had to take medicine |
| You had mastitis |
| You had sore or cracked nipples |
| You were told you had inverted nipples |
| You had difficulty attaching the baby |
| You had too much milk |
| Baby refused the breast |
| Baby had tongue tie |
| Breastfeeding was not convenient |
| You wanted to be able to leave the baby for several hours at a time |
| You had to return to work |
| The baby’s father didn’t want you to breastfeed |
| Other relative didn’t want you to breastfeed |
| You wanted to go on a diet |
| You needed somebody else to feed the baby |
| Someone else wanted to feed the baby |
| You wanted my body back to yourelf |
| You did not feel comfortable breastfeeding in public |

16. Out of these reasons, which was the most important?

17. If you are you are partially breastfeeding and partially formula feeding now, what are your current reasons for supplementing with formula? Please indicate all reasons that apply.

| Baby is not gaining enough weight (or losing weight) |
| --- |
| Baby is sick and cannot not breastfeed |
| You do not have enough supply |
| A health professional said I should for medical reasons |
| You are sick or taking medicine |
| You have mastitis |
| You have sore or cracked nipples |
| You were told you have inverted nipples |
| You are having difficulty with attaching the baby |
| You have too much milk |
| Baby refuses the breast |
| Baby has tongue tie |
| Breastfeeding is not convenient |
| You want to be able to leave the baby for several hours at a time |
| You have to return to work |
| The baby’s father doesn’t want you to breastfeed |
| Other relative doesn’t want you to breastfeed |
| You want to go on a diet |
| You need somebody else to feed the baby |
| Someone else wants to feed the baby |
| You want your body back to yourself |
| You do not feel comfortable breastfeeding in public |
| Other |

18. If you have ceased breastfeeding entirely what was the main reason?

| Baby was not gaining enough weight (or losing weight) |
| --- |
| You did not have enough milk supply |
| A health professional said you should for medical reasons |
| You had to stop to take medication |
| You had mastitis |
| You had sore or cracked nipples |
| Baby was not interested in breastfeeding |
| You were having difficulty with attaching the baby |
| You had to return to work |
| Other |

19. If you have ceased breastfeeding entirely what other reasons have contributed? (list as many as apply)

| Baby was not gaining enough weight (or losing weight) |
| --- |
| You did not have enough milk supply |
| A health professional said you should stop for medical reasons |
| You had to stop to take medication |
| You had mastitis |
| You had sore or cracked nipples |
| Baby was not interested in breastfeeding |
| You were having difficulty with attaching the baby |
| You had to return to work |
| Other |

20. Additional Participant Comments
